# FeverIQ - A Privacy-Preserving COVID-19 Symptom Tracker with 3.6 Million Reports

**DOI:** 10.1101/2020.09.23.20200006

**Authors:** Ankit Ranjan, Serena Li, Boyuan Chen, Karthik A. Jagadeesh, Alan Chiu, Jan Liphardt

## Abstract

Population-scale COVID-19 management benefits from timely and honest information from billions of people. Here, we provide a first report on the FeverIQ symptom tracker, a global effort to collect symptom and test data which has received more than 3.6 million submissions. Unlike other trackers, FeverIQ uses secure multiparty computation (SMC) to cryptographically guarantee user privacy while providing insights to scientists and public health efforts. We performed basic integrity checks of the FeverIQ dataset, such as by comparing it to other publicly released data. We then trained a linear classifier on diagnosis scores which were computed securely, without unprotected symptom data ever leaving a user’s phone or computer. FeverIQ is currently the world’s largest application of SMC in a health context, demonstrating the practicality of privacy-preserving analytics for population-scale digital health interventions.

## Introduction

COVID-19 is the first pandemic of the 21st century, with currently more than 25 million cumulative infections and 860,000 deaths.^1^ The SARS-CoV-2 virus spreads from person to person through physical contact and airborne droplets.^2^,3 Traditional strategies for managing pandemics combine testing and quarantines with contact tracing, and have proven effective for COVID-19^4^, but not all countries have been able to deploy these tools on necessary scales.^5^,6 The broad adoption of smartphones — currently exceeding 3.5 billion^7^ — suggests using them to augment classical epidemiological tools. In particular, smartphone data could help to map disease spread, detect at-risk individuals, facilitate contact tracing, and connect people with resources.

Current digital COVID-19 tools are focused on containment and research. For example, the Google/Apple Exposure Notification API allows users with smartphones to provide data for contact tracing and receive alerts about possible exposure. Symptom tracking projects such as the COVID Symptom Tracker^8^ and HowWeFeel^9^ are helping to uncover new symptoms and predict local trends. Mathematical modeling^10^ and practical evidence suggest that these tools can provide crucial information to individuals and public health systems.

Unfortunately, despite the clear utility of better and more timely data, efforts to deploy digital COVID-19 tools have struggled.^11^ There are numerous regulatory and political barriers to collecting and sharing health data, especially when it is identified. Equally problematic is that the relevant data include highly sensitive information such as with whom people interact, where they live, work and sleep, and medical test results for themselves and family members. People can be reluctant to divulge this information without a clear understanding of who will see the data, how long and where it will be stored, and what it will be used for.

We sought to reconcile the seemingly conflicting requirements of pandemic surveillance and user privacy. The role of privacy has not been broadly addressed in tracking projects, which collect participants’ symptoms, test results, and metadata. Especially for longitudinal studies, in which symptoms and geospatial data accumulate for each study participant, it can become possible to re-identify participants even when identifiers such as names and email addresses are scrubbed from the data.^12,13^ Unsurprisingly, lack of privacy reduces the honesty of self-reporting^14^ and may slow adoption in a global context. The World Health Organization has published guidance on the ethical considerations of contact-tracing and symptom tracking, with a key principle of data minimization to protect human rights.^15^ Overall, there is a pressing need for symptom tracking which maintain user privacy without compromising public health benefits.

## Approach and Results

### Privacy-Preserving Computation

Secure multiparty computation (SMC) denotes systems in which multiple agents are able to compute together without revealing their inputs. That capability is especially relevant to machine learning applications, where data security is of particular concern, either to protect data sources or the nature of the algorithms that are to be applied to the data. SMC protocols describe an online phase, during which parties jointly compute values, and offline phases, during which parties are capable of computing independently from others. The state of the art for SMC is the SPDZ protocol^16^, which increases offline computation and therefore the practicality of SMC for real-world applications. By implementing SPDZ, our computation requires at most five requests from client to server, which makes symptom tracking practical in a global, low resource context with unreliable internet and minimal phone hardware requirements.

### A Global, Private Symptom Tracker

We analyzed data from a global symptom tracking experiment run by Enya Inc., a confidential edge-computing company. Enya used secure multiparty computation to generate a COVID-19 symptom dataset with contributions from more than 3.6 million volunteers in 91 countries, including China, Vietnam, and the US. These data are available for academic research and public health efforts.

### Data Collection

Enya collected reports through the FeverIQ website (*https://map.feveriq.com*) and the FeverIQ web application integrated with the distributed Pi Social Network App.^17^ The survey consisted of a simple web form, which was intentionally minimal to easily load on edge devices. The survey asked for information about symptoms and the results of molecular SARS-CoV-2 testing. Upon data submission, participants were shown a global map with other people’s responses. To protect the participants’ privacy, no identifiable information was requested, collected, transmitted, or stored, and geolocation data were downsampled prior to leaving the user’s device.

### Characteristics of the study population

From 31 March 2020 to 4 September 2020, Enya collected 3,639,690 records from around the world, with the largest representations from China (49.05%), Vietnam (10.02%), and Egypt (4.86%) (see Figure 2). Around 300,000 records reported a diagnostic test result (50,804 positive, 251,144 negative).

### Secure Symptom Scoring

One challenge posed by SARS-CoV-2 is that it shares initial symptoms with other respiratory and immunological conditions. Therefore, the specific medical need is not only to detect some form of respiratory illness, but to discriminate among at least four competing alternatives: COVID-19, the common cold, the flu, and allergies. In the US, at least 1/5 of the population exhibits at least one respiratory symptom (e.g. fever, coughing, or sore throat) at any one time^18^, even outside of a typical flu season, and similar numbers hold for the rest of the world.^19^ The broad prevalence of non-COVID-19 respiratory illnesses with symptoms that overlap those of COVID-19 prompts the development of tools for more reliable discrimination of COVID-19 against a ‘noisy’ background of other illnesses with partially overlapping initial symptoms.

To address this need while cryptographically protecting the participant’s privacy, the web application used SMC to determine four scores designed to capture the similarity of the user’s symptoms to four preconfigured ‘diagnosis’ vectors: *COVID-19-base, COVID-19-neuro, cold*, and *flu*. During February and March, when new information about the presentation of COVID-19 was being published, diagnosis vectors were derived from multiple sources (publications in the medical literature, social media posts from front-line doctors in Wuhan, China and Italy, and statements from the CDC and the WHO) to represent two common manifestations of COVID-19 and the two main competing alternatives (Table 2). The *COVID-19-base* vector models ‘well-established’ symptoms (e.g. coughing, fever) whereas the *COVID-19-neuro* vector sought to capture emerging symptoms (e.g. hallucinations) which were just being reported. The *cold* and *flu* vectors represent symptoms of those well-understood respiratory illnesses, as found in the medical literature and in guidance from health authorities such as the CDC and the WHO.

### Secure Multiparty Computation

When a participant entered their symptoms, SMC was used to compute the inner product of their input vector with the four diagnosis vectors. This gave four inner product scores, typically ranging from zero to about 10. Specifically, Enya used two-party computation (2PC), a subproblem of SMC, with the patient and a server as each party. Matrix-vector multiplication was implemented with SPDZ, a secret-sharing-based approach to 2PC (Table 1). This approach depends on a pregenerated set of Beaver Triples^20^, which were created before the online phase by a third party. So-called oblivious transfer (OT) can be used to generate Beaver Triples^21^ but each scoring requires the generation of 64 triples, making it currently impractical to generate them with OT. Deploying Enya’s protocol requires the existence of a trusted third-party to generate triples, but that party does not need to be exposed to raw patient data or algorithm weights.

**Table 1.** SMC protocol for calculating the inner product scores.

| Step | Client | Server |
| --- | --- | --- |
| 1 | Secret share $X$ with the server | Secret share $M$ with the client |
| 2 | Compute and share $[[X]] - [[A]]$ and $[[M]] - [[B]]$ with the server | Compute and share $[[X]] - [[A]]$ and $[[M]] - [[B]]$ with the client |
| 3 | Reconstruct $X - A$ and $M - B$. Compute $[[Z]]^*$ and share with the server | Reconstruct $X - A$ and $M - B$. Compute $[[Z]]^*$ |
| 4 | | Reconstruct $Z$, a $n$ dimensional vector containing the diagnosis scores |

Later in the experiment, the inner products were supplemented with the Euclidean distances *D*(*X,Y*) between a user’s input vector and the diagnosis vectors, which define a space (Figure 1). To calculate the distances, the inner products ⟨*X,Y*⟩ were computed with SMC per Table 1 and, additionally, each party computed and shared their vector norms. The Euclidian distances were then obtained via 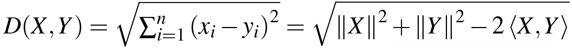.

**Figure 1.**
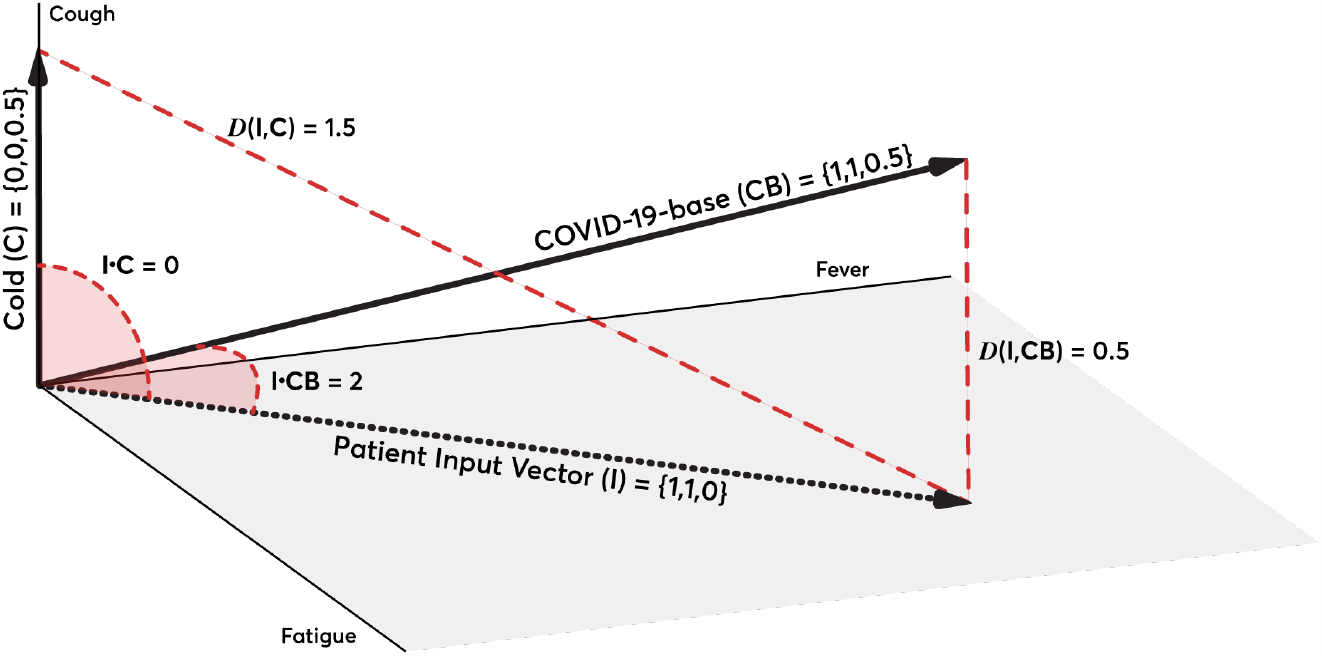
A simplified representation of an input vector from a person with fever and fatigue, but not cough. The *Cold* and *COVID-19-base* vectors represent the values of these diagnosis vectors in the dimensions of Fatigue, Fever, and Cough. The red dashed lines represent the Euclidean distances *D*(*X,Y*) and the red shaded areas denote the angles *θ* that are related to the inner products via 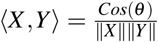.

**Figure 2.**
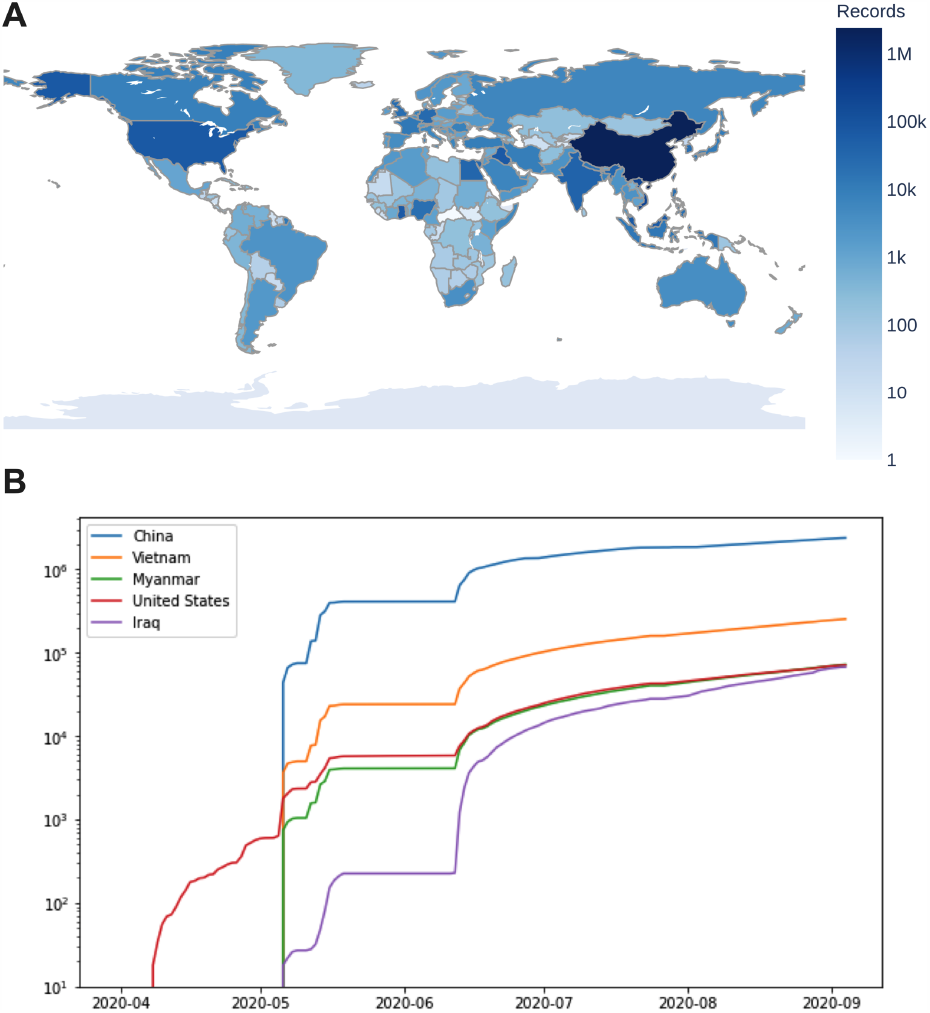
Global distribution of reports to FeverIQ in log scale. (**A**) shows countries reporting, with the largest number of reports from China and significant numbers from Vietnam and the US. (**B**) shows the cumulative reports over time from launch to 4 September 2020.

Mathematically, the inner product picks out the presence of specific symptoms without reducing the score when other, potentially unexpected symptoms are also present. By contrast, the Euclidean distances are useful to explore the entirety of the symptom space e.g. by clustering the participants’ scores and distances, as the starting point for refining or augmenting the diagnosis vectors. For example, if the *COVID-19-base* vector is deficient by lacking a COVID-19 symptom, then this would make itself known through a cluster of people in symptom space with high inner product scores but also large Euclidean distances, denoting additional information content not captured by a ‘trial’ *COVID-19-base* diagnosis vector.

### Data Analysis and Classification

We performed linear estimation on the inner product symptom scores for each of the four diagnosis vectors, by randomly dividing the data with complete input vectors and a reported test result into training and test sets (4:1 ratio). We evaluated four binary classifiers on the diagnosis scores: Linear Classifier (LC), Support Vector Classifier (SVC), Random Forest Classifier (RFC), and K-Nearest Neighbors (KNN), by comparing the the area under the curve of the receiver operator characteristic (AUC-ROC). The best performance was seen with LC and RFC, and we proceeded to assess the linear model for simplicity. To determine the best (combination of) input vectors for classification, we compared linear classifiers trained on each combination of the four vectors, again by comparing the AUC-ROC.

### Privately-collected data match established features of the disease

As we did not collect the date of molecular testing, we can not cross-check our data with public health records for regional positivity rates and trends. Especially for geographic regions with massive testing efforts but with only sporadic positive results, collecting the specific test date in concert with approximate geolocation could pose a potential risk to user privacy. Therefore test dates were not requested in the FeverIQ survey.

However, other features of the data can be used for quality checking. The FeverIQ data match several known characteristics of the disease and its spread, providing a first indication that the data contain relevant signal. First, the FeverIQ data measure a 25.3% asymptomatic positive rate for participants with a test result, which corresponds with other reports of 19 – 40%.^22^,23 Second, when cross-comparing the four diagnosis vectors, the two vectors which provided the best classification were the two COVID-19 vectors (*COVID-19-base* and *COVID-19-neuro*), as hoped, since those two vectors were derived from COVID-19 case reports and the medical literature, and therefore should have largest signal. Third, our reported ratios of positive tests to total tests (the ‘positivity rate’) coincide with publicly reported numbers for the five highest-reporting states in the US.

### Classification performs best with traditional and emerging diagnosis vectors

Our linear classifier achieved an AUC-ROC of 0.61 with the inner product diagnosis scores (Figure 3). Among all combinations of diagnosis vectors, the best performance was achieved with a combination of the *COVID-19-base* and *COVID-19-neuro* vectors. While we expected the related but negative diagnosis vectors for cold and flu to improve specificity, they had the opposite effect. When we run the linear classifier on all participants who reported symptoms and provided complete input vectors, we found 3.4% were predicted positive, in line with results seen in seroprevalence surveys, such as 1.5% in Santa Clara County^24^, although publicly available positivity data vary widely. The positive difference most likely reflects our selection for symptomatic reports.

**Figure 3.**
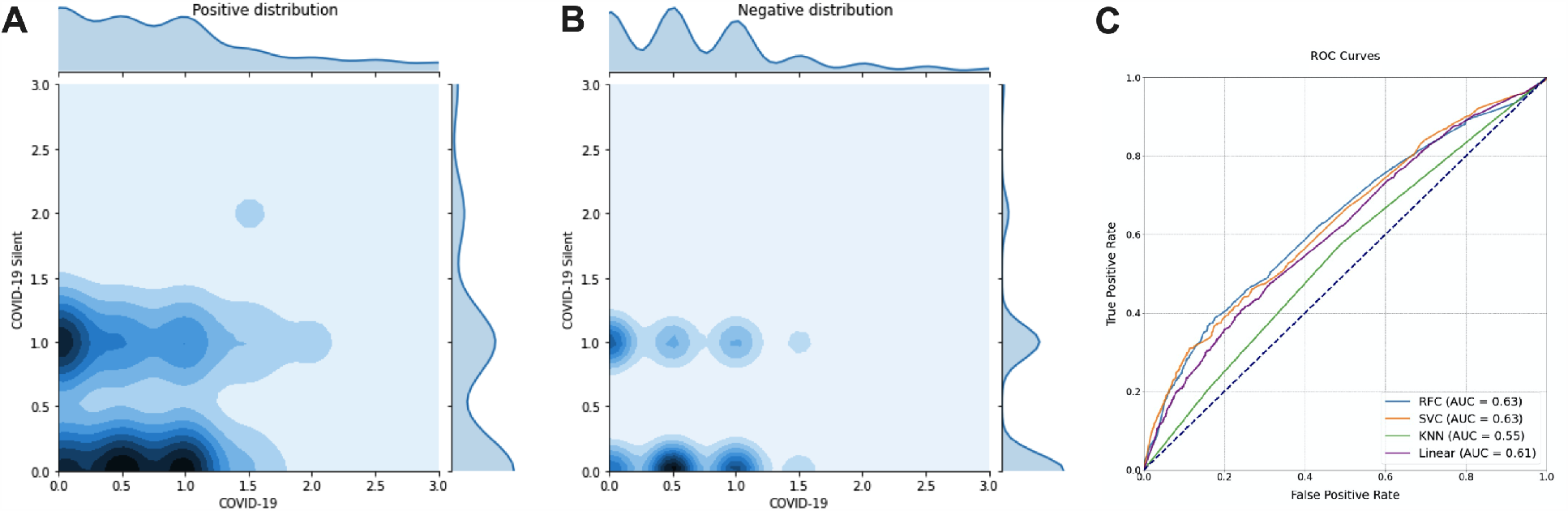
Classifier performance of different binary classifiers on inner product symptom scores. (**A**) shows the joint distribution of *COVID-19-base* and *COVID-19-neuro* symptom scores for positive results, and (**B**) shows this distribution for negative results. As expected, symptom scores are higher for positive records. (**C**) shows the ROC curves for four different binary classifiers on the symptom scores.

## Conclusions

### Limitations

One limitation of our work is the potential for false reporting and selection bias, by recruiting through a partner smartphone application. Honesty is seen to improve in private settings, so the use of privacy-preserving methods may reduce false-reporting. Another limitation is that, by design, our approach does not collect time or type of test for participants who report a test result. We cannot say for certain when symptoms arise in the course of the disease, and furthermore, this may affect the classification by including low-symptom or asymptomatic records in the positive test category for patients who have tested positive but since recovered. Finally, by using the inner product as the scoring metric in our SMC protocol, we emphasize the similarity between the input vectors and diagnosis vectors, and do not pay attention to symptoms missing in the profile, which may explain why the two best vectors for classification were COVID-19 diagnosis vectors. A future protocol may instead use the Hamming distance between the vectors to also reflect negative information.

Another limitation relates to iterative improvement of the diagnosis vectors. In the basic deployment, as implemented in the early weeks of the pandemic, was not possible to make post-hoc modifications (e.g. to the diagnosis vectors) to refine them or explore other ideas. Toward the end of the study, this limitation was partially alleviated by also computing Euclidean distances, which allows deficiencies in the diagnosis vectors to be discovered. For example, a cluster of people with high inner products *and* high Euclidean distances along one of the scoring axes indicates non-optimal settings for one or more of the diagnosis vector terms. Inability to train or refine classifiers is not a general limitation of privacy preserving analytics and there is a growing literature on techniques for updating model parameters in federated deep-learning applications without compromising security.^25^ However, when deploying privacy-preserving analytic tools to millions of users, care must be taken to contain costs, minimize data payload sizes and compute times, and ensure compatibility with rudimentary phone hardware without specialized cryptographic hardware.

### Outlook

Our work suggests that global-scale health needs can be balanced with user privacy. In particular, we are able to validate the diagnostic power of newly reported symptoms without needing to receive unprotected granular health information from participants. Furthermore, we are able to train a diagnostic classifier with private data that achieves comparable accuracy to classifiers trained on cleartext data. The best use case for this approach is when classifiers have been trained and validated on cleartext data, and then, one seeks to make these classifiers available to millions or billions of people without endangering their privacy. Privacy does not need to be compromised for symptom screening efforts in this pandemic and we provide a cryptographic and analytic framework able to handle millions of data submissions for risk scoring and disease surveillance.

## Data Availability

The FeverIQ dataset is available to researchers and public health officials. Please to request a copy of the data, providing your university or health system affiliation and a description of the intended research use. The full dataset is about 1GB in size. The analysis pipeline used for this report is available to modify as a Google Colab notebook.

## Acknowledgements

Enya Inc. gratefully acknowledges the Pi Network (*minepi*.*com*) members who submitted data about their symptoms and COVID-19 test reports. We thank Enya Inc. for generously making these data available to all qualified researchers. Analysis of the publicly available Enya data was performed at Stanford University and supported in part by the Bill & Melinda Gates Foundation.

## Enya Data Access Procedures

The FeverIQ dataset is available to researchers and public health officials. Please email *contact@enya*.*ai* to request a copy of the data, providing your university or health system affiliation and a description of the intended research use. The full dataset is about 1GB in size. The analysis pipeline used for this report is available to modify as a Google Colab notebook.

## Author contributions

A.R., B.C., K.J., A.C., and J.L. contributed to building FeverIQ. A.R., S.L., and J.L. contributed to data analysis and writing the paper. All authors reviewed the paper. K.J. is an advisor and J.L. is a co-founder of Enya.

## Supplementary Material

**Table 2.** The symptom weights for the four different disease conditions.

|  | COVID-19-base<sup>26</sup> | Cold<sup>27</sup> | Flu | COVID-19-neuro |
| --- | --- | --- | --- | --- |
| Fever | 1 | 0 | 1 | 0 |
| Fatigue | 0.5 | 0.5 | 1 | 0 |
| Cough | 1 | 0 | 1 | 0 |
| Sneezing | 0 | 1 | 0 | 0 |
| Aches | 0.5 | 1 | 1 | 0 |
| Runny Nose | 0 | 1 | 0.5 | 0 |
| Sore Throat | 0.5 | 1 | 0.5 | 0 |
| Diarrhea | 0.1 | 0 | 0.1 | 0 |
| Headaches | 0.5 | 0.1 | 1 | 0 |
| Short Breath | 0.5 | 0.1 | 0.1 | 0 |
| Anosmia | 1 | 0.1 | 0.1 | 0 |
| Allergies | 0 | 0 | 0 | 1 |
| Hallucinations | 0 | 0 | 0 | 1 |
| Vomiting | 0 | 0 | 0 | 1 |
| Blue Toes | 0 | 0 | 0 | 1 |
| Skipped Meals | 0 | 0 | 0 | 1 |

